# Identifying trends in SARS-CoV-2 RNA in wastewater to infer changing COVID-19 incidence: Effect of sampling frequency

**DOI:** 10.1101/2022.10.28.22281667

**Authors:** Elana M. G. Chan, Lauren C. Kennedy, Marlene K. Wolfe, Alexandria B. Boehm

## Abstract

SARS-CoV-2 RNA concentrations in wastewater solids and liquids are correlated with reported incident COVID-19 cases. Reporting of incident COVID-19 cases has changed dramatically with the availability of at-home antigen tests. Wastewater monitoring therefore represents an objective tool for continued monitoring of COVID-19 occurrence. One important use case for wastewater data is identifying when there are sustained changes or trends in SARS-CoV-2 RNA concentrations. Such information can be used to inform public health messaging, testing, and vaccine resources. However, there is limited research on best approaches for identifying trends in wastewater monitoring data. To fill this knowledge gap, we applied three trend analysis methods (relative strength index (RSI), percent change (PC), Mann-Kendall (MK) trend test) to daily measurements of SARS-CoV-2 RNA in wastewater solids from a wastewater treatment plant to characterize trends. Because daily measurements are not common for wastewater monitoring programs, we also conducted a downsampling analysis to determine the minimum sampling frequency necessary to capture the trends identified using the “gold standard” daily data. The PC and MK trend test appear to perform similarly and better than the RSI in terms of early warning signaling for increasing and decreasing trends, so we only considered the PC and MK trend test methods in the downsampling analysis. Using an acceptable sensitivity and specificity cutoff of 0.5, we found that a minimum of 4 samples/week and 5 samples/week is necessary to detect trends identified by daily sampling using the PC and MK trend test method, respectively. If a higher sensitivity and specificity is needed, then more samples per week would be needed. Public health officials can adopt these trend analysis approaches and sampling frequency recommendations to wastewater monitoring programs aimed at providing information on how incident COVID-19 cases are changing in the contributing communities.

## 1.0 Introduction

Public health departments have closely monitored cases of coronavirus disease 2019 (COVID-19) in their communities throughout the pandemic. Hospitals, healthcare providers, and laboratories are required to report incident laboratory-confirmed cases of COVID-19—hereafter referred to as incident clinical cases—to public health departments under state disease reporting laws [1]. This information allows health departments to track disease occurrence and may then be used to inform nonpharmaceutical interventions, such as mask mandates and social distancing, and education and outreach campaigns for testing and vaccines. Clinical test seeking behavior has changed dramatically with the availability of vaccines and at-home antigen tests [2], and results from the latter are not reported to health departments [2]. As a result, incident clinical case data may presently suffer from vast under-reporting relative to earlier in the pandemic and be less useful for tracking COVID-19 infections.

Wastewater-based epidemiology (WBE) uses concentrations of infectious disease markers in wastewater to track disease occurrence in the contributing community. It has been recommended by the World Health Organization (WHO) since 2003 for poliovirus monitoring in regions where polio is endemic [3]. The COVID-19 pandemic brought heightened attention to WBE which is currently in use globally for COVID-19 monitoring [4,5]. Specifically, RNA concentrations of severe acute respiratory syndrome coronavirus 2 (SARS-CoV-2) in primary wastewater settled solids are well-correlated with incident clinical COVID-19 cases in the same sewershed [6–8]. A recent study suggests that wastewater concentrations of SARS-CoV-2 RNA are also well-correlated with COVID-19 prevalence, which was estimated from randomized nasal swab sampling, more so than incident clinical cases because case data are prone to reporting biases such as underreporting of asymptomatic cases [9]. In addition, trends in wastewater concentrations of SARS-CoV-2 RNA were found to precede trends in incident clinical cases in communities [10–17]. WBE may therefore be a more reliable and objective tool than incident clinical case data for continued monitoring of COVID-19 because wastewater captures both symptomatic and asymptomatic individuals and does not depend on test seeking behavior or testing availability.

However, there is still uncertainty about how to appropriately interpret WBE data and use it to aid public health decision-making [18–20]. Accurately interpreting SARS-CoV-2 RNA concentrations as increasing, decreasing, or plateauing is important for guiding pandemic response efforts. Yet there has been limited work on how to actively monitor trends in epidemiology [21]. Common trend metrics (e.g., simple moving averages, rates of change) provide little attention to the statistical significance and stability of trends and can be misleading or confusing [17,21,22]. Standardized trend analysis methods that are robust and easily interpretable are needed to appropriately identify trends. Trend analysis of time-series data is conducted in other disciplines, such as finance [23], and could be adapted to interpret WBE data. Predicting the stock market in real time is desirable to traders similar to how predicting the course of disease occurrence is useful to public health decision makers, and both price data and WBE data have stochastic elements [23]. One goal of this study was to test different trend analysis methods for application to WBE.

When analyzing time-series data for trends, a large number of observations provides greater statistical power [24], and high frequency (e.g., daily) data has previously been identified as ideal for WBE to most accurately identify trends [25]. However, daily sampling and processing of wastewater can be challenging to implement [18,20], and many facilities across the United States only collect samples once per week [26,27]. Previous studies found that sampling wastewater at least twice per week is needed to detect correlations between wastewater SARS-CoV-2 RNA concentrations and incident clinical cases [6,28,29], but these studies collected data over a limited duration of time (at most six months) or did not include daily data in the analysis. In addition, the authors examined the correlation between wastewater SARS-CoV-2 RNA concentrations and incident clinical cases to come to their conclusions. Research is needed to investigate how frequently wastewater should be sampled if the goal of WBE is to correctly identify trends in wastewater concentrations of SARS-CoV-2 RNA.

In this study, we used daily measurements of SARS-CoV-2 RNA in wastewater from a wastewater treatment plant between November 2020 and September 2022 to (i) compare three different trend analysis methods for characterizing trends in wastewater SARS-CoV-2 RNA concentrations and (ii) evaluate the performance of each method using data sampled at a lower frequency than one time per day. This dataset is ideal because it spans nearly two years of the pandemic, including three major waves (Delta, BA.1 Omicron, and BA.2 + BA.5 Omicron). These waves differ in magnitude and shape, so this dataset can be used to examine how trend analysis methods respond across a variety of disease dynamics. This dataset is also ideal for a downsampling analysis because it can be downsampled to all possible sampling frequencies (1 sample/week to 6 samples/week). We identify robust trend analysis methods and recommend sampling frequencies that can be used by WBE programs to provide insight about the disease burden of COVID-19 in the contributing population.

## 2.0 Methods

### 2.1. Wastewater data

The San José-Santa Clara Regional Wastewater Facility serves 1.4 million residents and over 17,000 businesses throughout Silicon Valley (Fig 1 in S1 Text). The wastewater treatment plant is the largest advanced wastewater treatment plant in the western United States and treats 110 million gallons of wastewater per day on average with a capacity of 167 million gallons per day [30].

**Fig 1.**
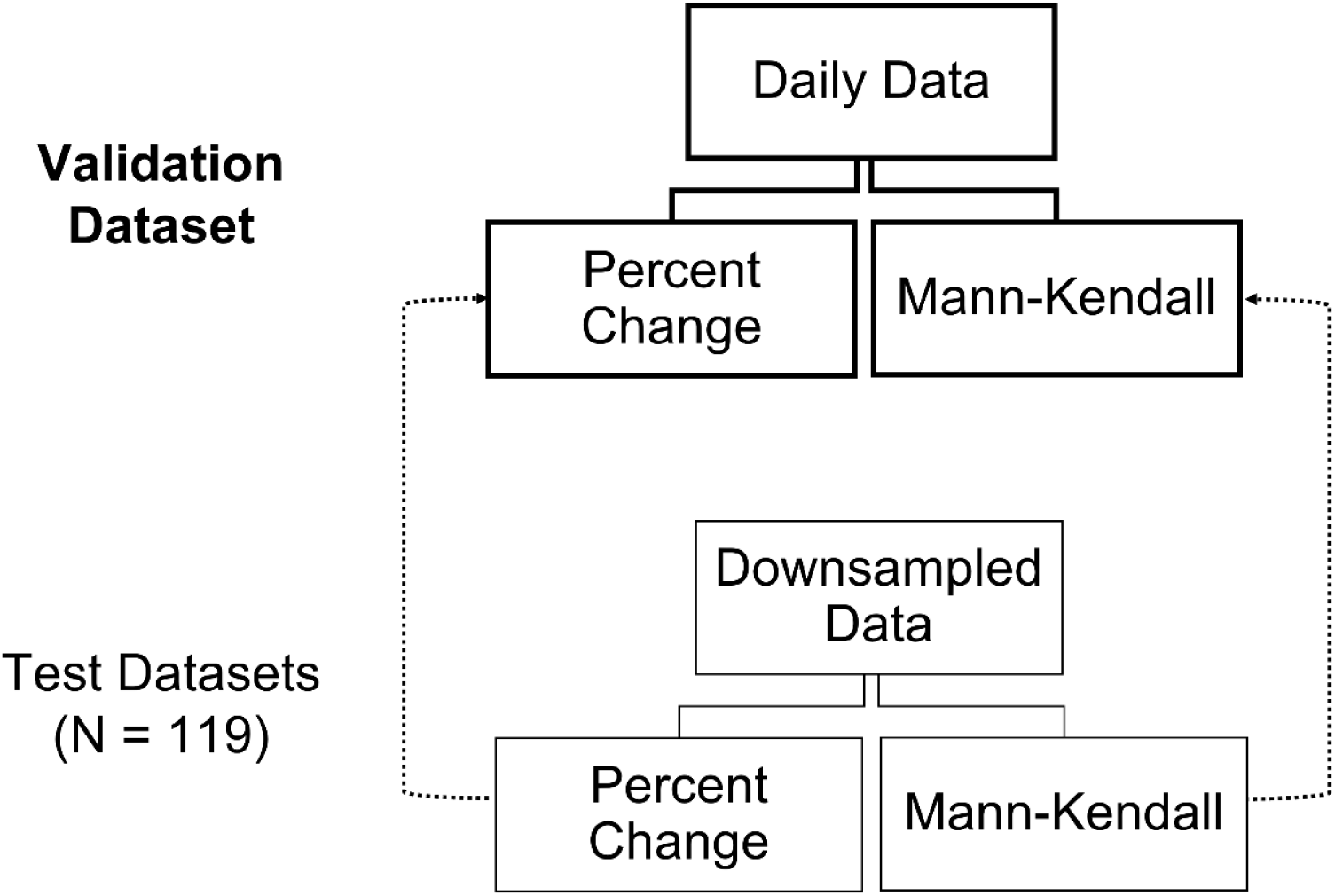
Process for sensitivity and specificity calculations. Each method was (i) applied to the daily dataset and (ii) applied to each downsampled dataset (N = 119). Then for each method separately, the trend analysis results from each downsampled dataset were validated with the trend analysis results from the daily dataset

Daily sampling from the wastewater facility began on November 15, 2020, and we used data through September 15, 2022, for this study (n = 670 days). Sampling and processing details, including quality assurance and quality control metrics, are registered in protocols.io [31–33] and have been described previously by Kim et al. [34] and Wolfe et al. [35]. For this analysis, we used concentrations of the SARS-CoV-2 RNA N gene in gene copies per gram of dry weight normalized by concentrations of pepper mild mottle virus (PMMoV) in gene copies per gram of dry weight (N/PMMoV). PMMoV is used as a marker of wastewater fecal strength [36,37]. There were no non-detects for either the N gene or PMMoV in the dataset. Data between November 15, 2020, and March 31, 2021, have been previously published by Wolfe et al. [35] and are publicly available through the Stanford Digital Repository (https://doi.org/10.25740/bx987vn9177) [38]. Data between January 1, 2022, and April 12, 2022, have been previously published by Boehm et al. [39] and are publicly available through the Stanford Digital Repository (https://doi.org/10.25740/cf848zx9249) [40]. The remaining data (April 1, 2021–December 31, 2021, and April 13, 2022–September 15, 2022) have not been previously published. All data used in this study are available publicly through the Stanford Digital Repository (https://doi.org/10.25740/yg713sw8276) [41]. Since the pre-analytical and analytical methods used for measuring the SARS-CoV-2 RNA N gene and PMMoV are registered and previously described in peer-reviewed publications, they are not repeated herein.

### 2.2. Trend analysis methods

We considered three metrics for identifying trends in N/PMMoV over time: relative strength index (RSI), percent change (PC), and the Mann-Kendall (MK) trend test. RSI is a common technical indicator used in finance for trend analysis [42], PC is used by the United States Centers for Disease Control (CDC) to report trends for the National Wastewater Surveillance System (NWSS) [43], and the MK trend test is a statistical test used to evaluate the existence of a monotonic trend in time-series data [44–46]. As described below, the RSI informs about the stability of a trend and PC and the MK trend test inform about the statistical significance of a trend. All calculations were conducted in R (version 4.1.3) [47]. Data and R code are available publicly through the Stanford Digital Repository (https://doi.org/10.25740/yg713sw8276) [41].

The RSI is a momentum indicator used in technical trading systems that measures the speed and direction of price changes over a specified time period [42]. The RSI is calculated using the relative strength (RS) which is the ratio of the average increase (gain) and decrease (loss) of closing prices over the look-back period (typically 14 days) (Equation 1) [42]. Refer to the S1 Text for further details. The RSI ranges from 0 to 100; values above 70 and below 30 signify an overbought and oversold market, respectively [42]. Here we calculated the RSI of the 7-day right-aligned moving average (MA) of N/PMMoV using a 14-day look-back period. Using the raw, or unaltered, N/PMMoV data resulted in a highly variable RSI that was unlikely to be useful (Fig 2 in S1 Text). Previous work that applied the RSI to incident clinical case data early in the pandemic also calculated the RSI of smoothed input data [21]. Based on previously published work [21,42,48], we interpreted RSI values above 70, 80, and 90 as upward, likely upward, and very likely upward trends, respectively. We interpreted RSI values below 30, 20, and 10 as downward, likely downward, and very likely downward trends, respectively (Table 1).

**Table 1.** Trend classification criteria for each trend analysis method.

| **Trend Classification** | **Relative Strength Index** | **Percent Change** | **Mann-Kendall Test** |
| --- | --- | --- | --- |
| **Very likely upward** | RSI ≥ 90 | 99% lower CI $>$ 0 | $p \leq 0.01, \tau > 0$ |
| **Likely upward** | $90 > \text{RSI} \geq 80$ | 95% lower CI $>$ 0 | $0.01 < p \leq 0.05, \tau > 0$ |
| **Upward** | $80 > \text{RSI} \geq 70$ | 90% lower CI $>$ 0 | $0.05 < p \leq 0.1, \tau > 0$ |
| **No Trend** | $70 > \text{RSI} > 30$ | all CIs intersect 0 | $p > 0.1$ |
| **Downward** | $30 \geq \text{RSI} > 20$ | 90% upper CI $<$ 0 | $p \leq 0.01, \tau < 0$ |
| **Likely downward** | $20 \geq \text{RSI} > 10$ | 95% upper CI $<$ 0 | $0.01 < p \leq 0.05, \tau < 0$ |
| **Very likely downward** | $10 \geq \text{RSI}$ | 99% upper CI $<$ 0 | $0.05 < p \leq 0.1, \tau < 0$ |

CI = Confidence Interval
RSI = Relative Strength Index
PC = Percent Change
MK = Mann-Kendall

**Fig 2.**
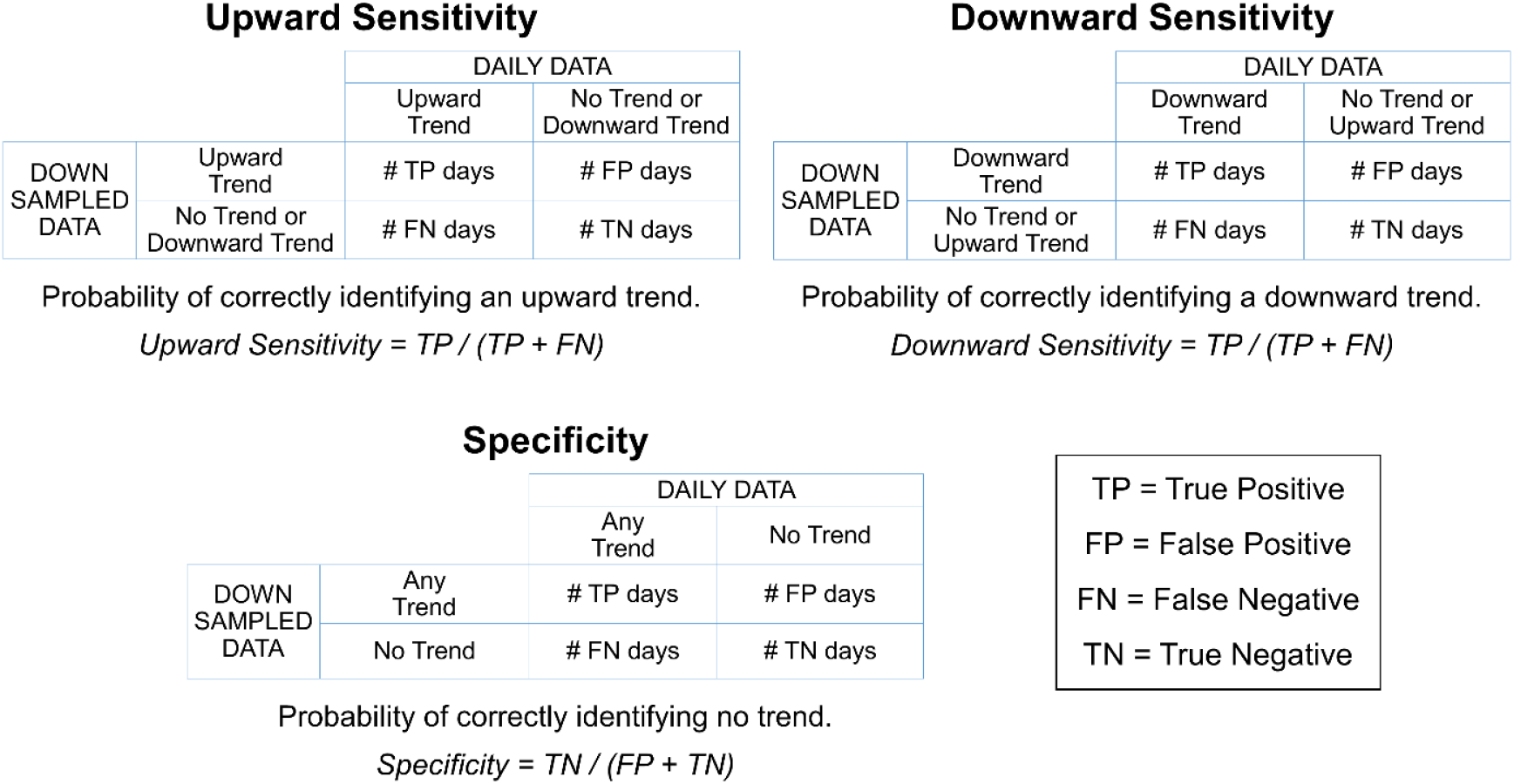
Definitions and equations used to calculate upward sensitivity, downward sensitivity, and specificity. Here an upward trend includes “very likely upward”, “likely upward”, and “upward” trends and a downward trend includes “very likely downward”, “likely downward”, and “downward” trends.

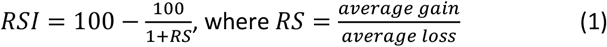

The second trend analysis method we investigated was PC. Specifically, we used the formula that the United States CDC uses to calculate trends in wastewater SARS-CoV-2 RNA levels for the NWSS (Equation 2). The slope is calculated from a least-squares linear regression of log10-transformed values of the raw N/PMMoV data versus day because wastewater SARS-CoV-2 RNA concentrations are log-normally distributed [43]. The CDC uses a 15-day look-back period to calculate the slope [43]; here we used a 14-day look-back period for consistency and because 14 days represents an integer multiple of one week. For each slope estimate, we extracted the 90%, 95%, and 99% confidence intervals (CIs) to calculate the 90%, 95%, and 99% CIs for each PC estimate. We interpreted PC values greater than 0 for the 90%, 95%, and 99% upper CIs as upward, likely upward, and very likely upward trends, respectively. We interpreted PC values less than 0 for the 90%, 95%, and 99% lower CIs as downward, likely downward, and very likely downward trends, respectively (Table 1).

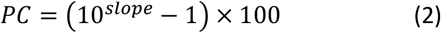

Lastly, the MK trend test is a nonparametric test that evaluates whether there is a monotonic trend in a time-series dataset [44–46]. Previous work that applied the MK trend test to incident clinical COVID-19 case data explored 5, 7, and 14-day look-back periods to identify statistically significant trends in COVID-19 case rates [22]. Here we applied the MK trend test to raw N/PMMoV data and used a 14-day look-back period to be consistent with the look-back periods used for RSI and PC. Log-transformation of the raw N/PMMoV values was not necessary because the MK trend test is nonparametric. We interpreted positive values of the test statistic, tau (τ), with p ≤ 0.1, p ≤ 0.05, and p ≤ 0.01 as upward, likely upward, and very likely upward trends, respectively. We interpreted negative τ values with p ≤ 0.1, p ≤ 0.05, and p ≤ 0.01 as downward, likely downward, and very likely downward trends, respectively (Table 1).

### 2.3. Application of trend analysis methods to wastewater data

First, we applied each trend analysis method to daily N/PMMoV measurements. Heatmaps were generated to visualize the trend classifications in Table 1 over the entire analysis period (November 15, 2020–September 15, 2022). Moreover, we created separate heatmaps for three major COVID-19 waves that occurred during our analysis period. Here, a wave is defined as a substantial rise and eventual decline in N/PMMoV concentrations caused by one or more SARS-CoV-2 variants as described by Wolfe et al. [49] and Boehm et al. [39] for this wastewater treatment plant. During the analysis period, the three waves that occurred were caused by the following SARS-CoV-2 variants: Delta, BA.1 Omicron, and BA.2 + BA.4 + BA.5 Omicron. Note that the BA.4 Omicron mutation was very rare during the latter wave [50,51], so we will herein refer to the BA.2 + BA.4 + BA.5 Omicron wave only as the BA.2 + BA.5 Omicron wave. The separate heatmaps allowed us to evaluate the performance, which is subjectively and qualitatively described, of the trend analysis methods during different phases of the pandemic.

Next, we downsampled the daily dataset and reapplied the PC and MK trend test methods. We did not consider the RSI method in the downsampling analysis because the RSI performed the poorest in the trend analysis using daily data (presented below). We created a downsampled dataset for all possible sampling combinations encompassing sampling frequencies between 2 samples/week and 6 samples/week (N = 119) (Table 2). Note that it is not possible to calculate PC or conduct the MK trend test using a 14-day look-back period and 1 sample/week frequency because these methods require at least three measurements, so we did not consider any 1 sample/week sampling combinations in our analysis. The total number of possible downsampling combinations *N* was calculated using Equation 3 where *f* is the number of samples collected per week (*f* = 2 for 2 samples/week, *f* = 3 for 3 samples/week, etc.). Seven was used in the numerator of the binomial coefficient because there are seven days in a week. For example, there are 21 unique pairs of days for a 2 samples/week sampling frequency. We did not impute missing N/PMMoV values for days designated as non-sampling days during the downsampling process (i.e., we kept missing data as missing in the downsampled datasets).

**Table 2.** Number of sampling combinations for each downsampling frequency.

| Downsampling Frequency | Number of Sampling Combinations |
| --- | --- |
| 2 samples/week | 21 |
| 3 samples/week | 35 |
| 4 samples/week | 35 |
| 5 samples/week | 21 |
| 6 samples/week | 7 |

Number of sampling combinations = 7 choose $f$ , where $f$ is the number of samples collected per week.

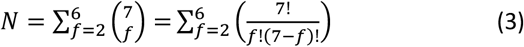

To evaluate the impact of using downsampled data for the PC and MK trend test methods, we calculated the sensitivity and specificity for each trend analysis method for each downsampled dataset using the daily dataset as the validation dataset for each method (Fig 1). Sensitivity was defined as the probability of a downsampled dataset to correctly identify when a trend was identified using the daily dataset (i.e., correctly identify true positives); specificity was defined as the probability of a downsampled dataset to correctly identify when no trend was identified using the daily dataset (i.e., correctly identify true negatives). Because the trend methods classify two distinct trend types (upward versus downward), we calculated upward sensitivity and downward sensitivity separately (Fig 2). For the sensitivity calculations, an upward trend includes “very likely upward”, “likely upward”, and “upward” trend classifications and a downward trend includes “very likely downward”, “likely downward”, and “downward” trend classifications from Table 1.

## 3.0 Results

We used three different trend analysis methods to characterize trends in wastewater SARS-CoV-2 RNA concentrations: the relative strength index (RSI) used in finance [42], the percent change (PC) method used by the United States CDC [43], and the nonparametric Mann-Kendall (MK) trend test [44–46]. The SARS-CoV-2 RNA dataset contained visual periods of increase, decrease, and stability in N/PMMoV concentrations, allowing us to evaluate the trend analysis methods throughout different COVID-19 waves. We then reapplied the PC and MK trend test methods to downsampled data to investigate the impact of using data sampled at a frequency of less than once per day.

### 3.1. Trend analysis methods applied to daily data

Using the daily dataset, wastewater SARS-CoV-2 RNA trends were classified each day as increasing, decreasing, or stable using three trend analysis methods (RSI, PC, MK trend test) (Fig 3A). Excluding the wave at the start of the dataset, three major waves occurred during our analysis period. The Delta wave was a relatively small wave (roughly identified as starting the second week of June 2021 and ending the first week of September 2021) (Fig 3B), the BA.1 Omicron wave was a relatively large wave with rapid onset (roughly identified as starting the third week of December 2021 and ending the fourth week of January 2022) (Fig 3C), and the BA.2 + BA.5 Omicron wave was a relatively large wave with gradual onset (roughly identified as starting the first week of April 2022 and ending the second week of September 2022) (Fig 3D). Daily N/PMMoV measurements with a 7-day moving average are shown above each heatmap for reference.

**Fig 3.**
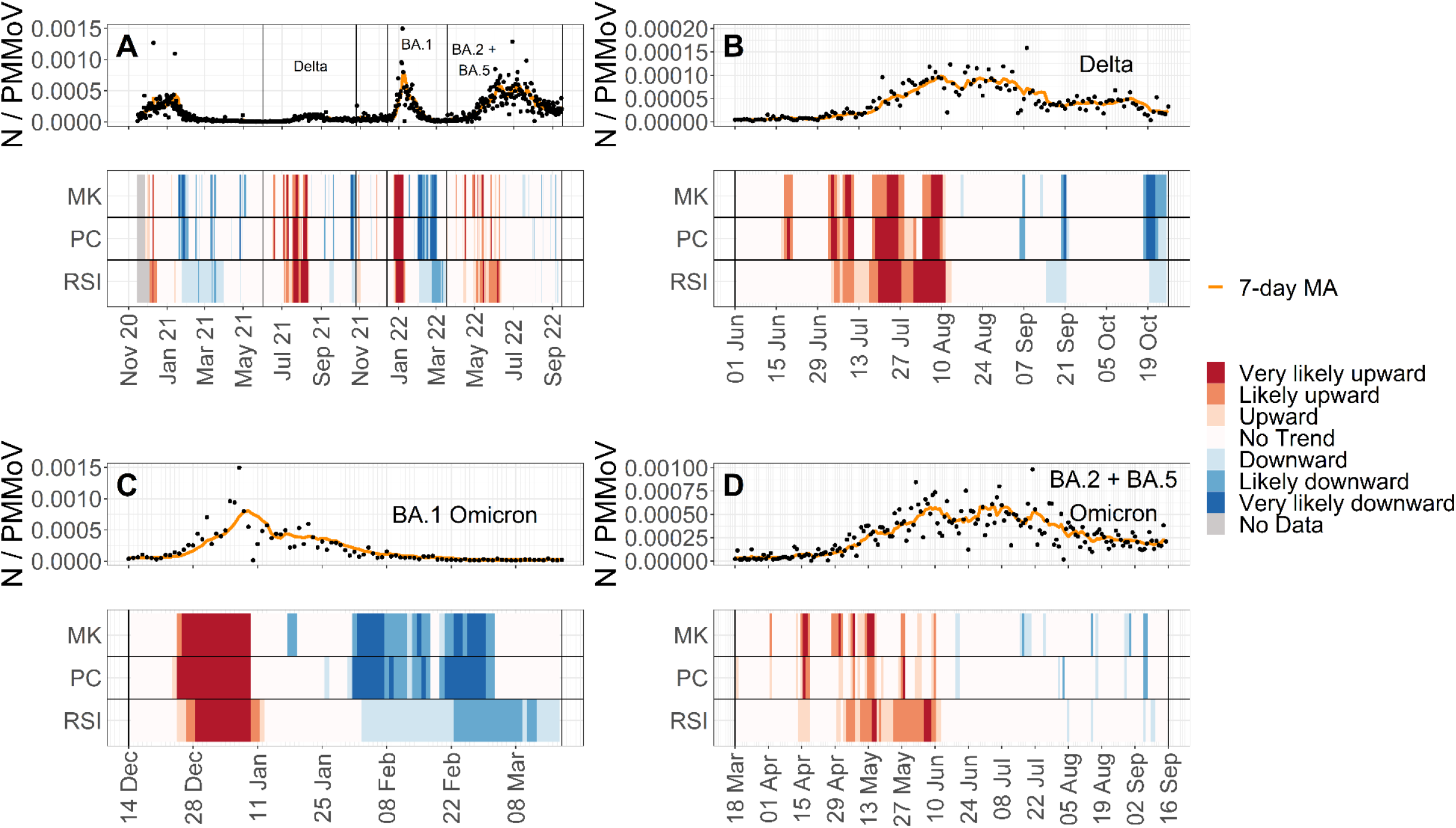
N/PMMoV with a 7-day moving average (MA) and corresponding trend classifications from relative strength index (RSI), percent change (PC), and the Mann-Kendall (MK) trend test using daily data. (A) Entire analysis period, (B) Delta wave, (C) BA.1 Omicron wave, and (D) BA.2 + BA.5 Omicron wave. Dates in panel A are given as “month year”; dates in all other panels are given as “day month”. “No Data” indicates that not enough N/PMMoV measurements were available yet to conduct the trend analysis method.

All three methods identified periods of increase, decrease, and no change in the wastewater concentrations. The PC and MK trend test methods identified an increasing trend sooner than the RSI at the start of the Delta and BA.2 + BA.5 Omicron wave. Specifically, PC and the MK trend test identified an increasing trend 16 and 17 days, respectively, before the RSI at the start of the Delta wave and 26 and 12 days, respectively, before the RSI at the start of the BA.2 + BA.5 Omicron wave. All three methods first identified an increasing trend at the start of the BA.1 Omicron wave within one day of each other. The PC and MK trend test also identified decreasing trends at the end of the waves prior to the RSI, in most cases over a week in advance of the RSI. Given the apparent tendency of the PC and MK trend test methods to identify changing conditions in advance of the RSI and that the PC and MK trend test are more rigorous than the RSI because they require some measure of statistical significance and hypothesis testing, the RSI method was not further included in the downsampling analysis portion of the study.

### 3.2. Downsampling analysis

For each downsampled dataset, wastewater SARS-CoV-2 RNA trends were classified each day as increasing, decreasing, or stable using the PC and MK trend test methods. Then for each method, the trend analysis results from each downsampled dataset were validated using the trend analysis results from the daily dataset by calculating upward sensitivity (ability to correctly identify an upward trend), downward sensitivity (ability to correctly identify a downward trend), and specificity (ability to correctly identify no trend). For both PC and the MK trend tests, upward and downward sensitivities were poor at low sampling frequencies but improved as sampling frequency increased. Using a median sensitivity better than chance (0.5) as an acceptable cutoff, PC achieved acceptable upward and downward sensitivity with at least 4 samples/week (Fig 4, Table 3). The MK trend test achieved acceptable upward and downward sensitivity with at least 5 samples/week (Fig 4, Table 3). Specificity was similar—and very high—at all sampling frequencies for both methods. A key limitation, however, is that the sensitivity and specificity values cannot be compared between methods because we validated each method with itself so their validation datasets differ (Fig 1)

**Table 3.** Upward sensitivity, downward sensitivity, and specificity of downsampled data for percent change and Mann-Kendall methods.

| **PERCENT CHANGE (PC)** |  |  |  |  |  |
| --- | --- | --- | --- | --- | --- |
| **Sampling Frequency**<br>**(sampling combinations)** | **2 samples/week**<br>**(n = 21)** | **3 samples/week**<br>**(n = 35)** | **4 samples/week**<br>**(n = 35)** | **5 samples/week**<br>**(n = 21)** | **6 samples/week**<br>**(n = 7)** |
| **Sensitivity**<br>**(upward trend)** | 0.28<br>(0.13) | 0.46<br>(0.08) | 0.60<br>(0.07) | 0.72<br>(0.06) | 0.80<br>(0.03) |
| **Sensitivity**<br>**(downward trend)** | 0.21<br>(0.09) | 0.38<br>(0.12) | 0.52<br>(0.11) | 0.67<br>(0.08) | 0.79<br>(0.06) |
| **Specificity**<br>**(no trend)** | 0.91<br>(0.03) | 0.90<br>(0.02) | 0.92<br>(0.02) | 0.93<br>(0.02) | 0.96<br>(0.01) |
| **MANN-KENDALL (MK) TREND TEST** |  |  |  |  |  |
| **Sampling Frequency**<br>**(sampling combinations)** | **2 samples/week**<br>**(n = 21)** | **3 samples/week**<br>**(n = 35)** | **4 samples/week**<br>**(n = 35)** | **5 samples/week**<br>**(n = 21)** | **6 samples/week**<br>**(n = 7)** |
| **Sensitivity**<br>**(upward trend)** | 0.23<br>(0.10) | 0.29<br>(0.11) | 0.50<br>(0.10) | 0.71<br>(0.07) | 0.87<br>(0.04) |
| **Sensitivity**<br>**(downward trend)** | 0.18<br>(0.10) | 0.26<br>(0.10) | 0.44<br>(0.11) | 0.56<br>(0.10) | 0.79<br>(0.07) |
| **Specificity**<br>**(no trend)** | 0.92<br>(0.02) | 0.95<br>(0.02) | 0.95<br>(0.02) | 0.95<br>(0.02) | 0.95<br>(0.02) |

Median is provided with standard deviation (SD) in parentheses.

**Fig 4.**
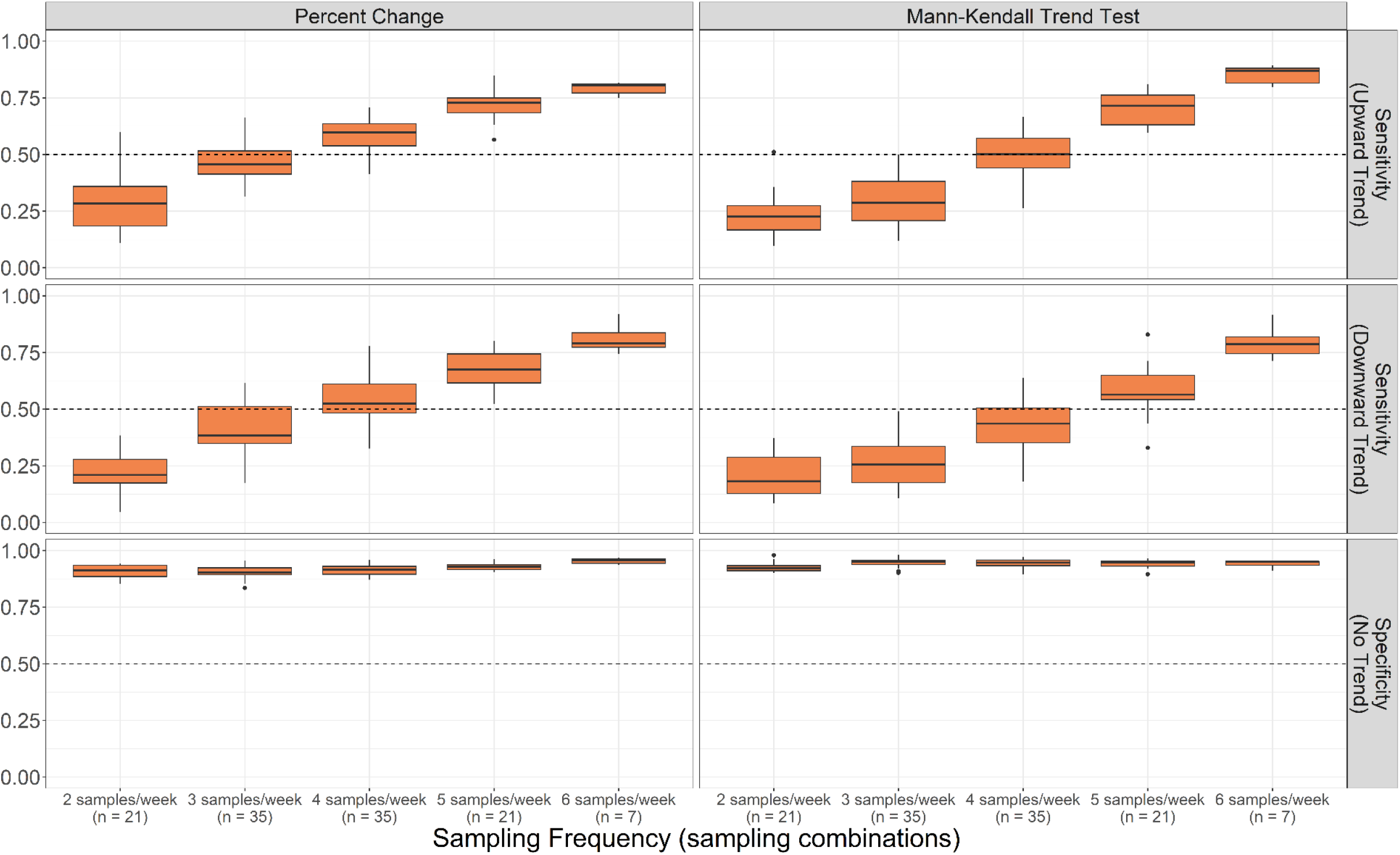
Upward sensitivity, downward sensitivity, and specificity of each downsampled dataset for the percent change and Mann-Kendall trend test methods. Results are aggregated by sampling frequency.

## 4.0 Discussion

We applied three different trend analysis methods to daily measurements of SARS-CoV-2 RNA from the San Jose sewershed between November 15, 2020, and September 15, 2022. We then created downsampled datasets representing all possible sampling combinations and reapplied the PC and MK trend test methods to each downsampled dataset. We evaluated the performance of each method conducted with downsampled data by calculating sensitivity and specificity, using the results from each method conducted with daily data as a validation dataset.

The PC and MK trend test methods appeared to provide more early warning signaling of upward and downward trends compared to the RSI when conducted using daily data. The PC method is currently used at the national level by the United States CDC for the NWSS [43], and a trend analysis of incident clinical COVID-19 cases found that the MK trend test was both intuitive and accurate [22]. These two methods are also more rigorous than the RSI method because they require some measure of statistical significance and hypothesis testing to classify trends (Table 1). It is important to note, however, that we could not quantitatively evaluate the accuracy of each method because there is no true validation dataset to identify when upward, downward, or no trends in SARS-CoV-2 RNA concentrations truly did occur. Even a potential validation dataset based on clinical case data is subject to biases from test seeking behavior and availability or delays due to reporting. We only had the benefit of hindsight to qualitatively compare the three methods which, in practice, would be applied to incoming wastewater data in real time.

The trend analysis methods presented here report trends based on the likelihood of a trend. Alternatively, health departments could report trends based on the duration over which a trend occurs instead of the likelihood of a trend. A study by Holst et al. classified trends in wastewater SARS-CoV-2 RNA concentrations as sustained increase, increase, plateau, decrease, or sustained decrease [25]. Sustained versus unsustained trends were differentiated based on the number of samples over which a statistically significant slope occurred (five samples for sustained trends and three samples for unsustained trends) [25]. Although beyond the scope of this study, the criteria used to classify trends could be amended to consider the duration over which trends occur.

Our downsampling analysis suggests that a minimum sampling frequency of 4 samples/week and 5 samples/week is necessary to detect trends that were identified by daily sampling using the PC and MK trend test, respectively. This is based on an acceptable median sensitivity and specificity better than chance (0.5). Previous studies that examined correlations between wastewater SARS-CoV-2 RNA concentrations and incident clinical COVID-19 cases found that collecting a minimum of 2 samples/week was necessary to detect significant correlations [6,28,29]. Our results suggest that collecting only 2 samples/week is not sufficient to detect trends identified by daily sampling using a 14-day look-back period when wastewater SARS-CoV-2 RNA concentrations are analyzed independently from incident clinical cases. We also conducted the downsampling analysis using a 21-day look-back period for each method and found similar results (i.e., a minimum sampling frequency of 3 samples/week and 4 samples/week is necessary to detect trends identified by daily sampling using PC and the MK trend test, respectively).

The PC and MK trend test methods appear to report similar trends for most days in the study period, but the PC method requires fewer samples per week than the MK trend test method to detect trends identified in the daily data with acceptable sensitivity and specificity. Therefore, the PC method may be preferred by WBE programs with potential budget constraints. According to our analysis, the PC method can still provide sufficient trend information using 4 samples/week. Further analysis could investigate which four-day sampling combinations tend to have an upward sensitivity, downward sensitivity, and specificity all above 0.5 (i.e., days clustered together versus days evenly spaced throughout the week). Additionally, WBE programs could adopt an adaptive approach such that sampling frequency is increased when there is a strong need to know the current trend (e.g., when COVID-19 incidence is suspected to be high or increasing).

Our analysis has some limitations. First, we applied the three trend analysis methods to wastewater data from one sewershed. The sewershed is large, so it is possible that the trend analysis methods may perform differently or that the downsampling results may differ in a smaller sewershed with more day-to-day variability in SARS-CoV-2 RNA concentrations. Furthermore, there were no days in our analysis period in which the N gene was below the limit of detection. It remains unclear how each trend analysis method performs or how the downsampling results may be affected by brief or prolonged periods in which the N gene concentration is below detection levels.

Second, our trend methods only report trend directions; they do not describe the magnitude of trends. For example, a small percent increase (e.g., <1%) would be classified as an upward trend if the lower CI is above 0% and a large percent increase (e.g., >10%) would be classified as no trend if the lower CI is below 0% according to our trend classification criteria—even though the magnitude of the latter trend is larger. The CDC does not consider statistical significance and only considers magnitude when classifying trends using the PC approach. Specifically, the CDC classifies trends into five categories based on the magnitude of the PC estimate over the last 15 days: large decrease (−100% or less), decrease (−99% to - 10%), stable (−9% to 9%), increase (10% to 99%), and large increase (100% or more) [52]. However, categorizing PC trends based on magnitude alone can be misleading when wastewater SARS-CoV-2 concentrations are low or around the limit of detection [17]. If public health departments want to report both the significance and magnitude of trends, the criteria in Table 1 could be modified such that the value of the PC or tau estimate must be both significant and above or below a certain threshold. The RSI only describes trend stability; it does not have an associated statistical significance and does not provide insight about the magnitude of a trend.

Third, trend analysis methods only describe whether wastewater SARS-CoV-2 RNA concentrations are increasing, decreasing, or remaining stable. Trend analysis methods do not describe the abundance of SARS-CoV-2 RNA and whether the quantity is high, medium, or low. Trends could be reported alongside thresholds to better contextualize the current state of the pandemic. High, medium, and low thresholds could be constructed based on relative concentrations (e.g., percentage of the maximum concentration during a recent wave or past time frame) or absolute concentrations. Future work is needed to determine how best to construct thresholds and which cutoff values to use to differentiate high versus medium versus low SARS-CoV-2 RNA quantities.

Notwithstanding these limitations, our analysis is highlighted by several strengths. Our analysis period spans nearly two years, so we were able to explore how each trend analysis method performed in response to three distinct COVID-19 waves. In particular, two of our trend analysis approaches (PC and the MK trend test) addressed the statistical significance of trends, which is currently not typically reported alongside trends [22], because we categorized trends for these methods based on statistical significance. In contrast, the United States CDC categorizes trends on the NWSS dashboard based only on trend magnitude and not whether the observed trend is statistically significant [52]. Additionally, our dataset contained daily SARS-CoV-2 RNA measurements which was ideal for a downsampling analysis. We were able to downsample this dataset to test all 119 possible sampling combinations ranging from 2 samples/week to 6 samples/week which allowed us to observe the full range of sensitivity and specificity values and, in turn, improve confidence in our sampling frequency recommendations.

During the COVID-19 pandemic, it has been demonstrated that WBE is a useful tool for public health monitoring, and the per capita cost of wastewater sampling is significantly lower than the per capita cost of individual clinical testing [53]. WBE has also been shown to be useful for monitoring the occurrence of other diseases [54–56]. We expect the analyses performed herein to be useful for interpreting wastewater monitoring data for COVID-19 and other infectious disease markers.

## 5.0 Conclusion

We compared three trend analysis methods for characterizing trends in SARS-CoV-2 RNA concentrations in wastewater. Based on daily data from the San Jose wastewater treatment plant, the PC and MK trend test appear to perform similarly and better than the RSI in terms of early warning signaling. Additionally, both the PC and MK trend test are inference-based methods so can be used to classify trends in a standard, statistically sound manner. When using the PC method to classify trends, our downsampling analysis suggests that a minimum sampling frequency of 4 samples/week is necessary to detect trends identified by daily sampling (5 samples/week using the MK trend test method). WBE programs can adopt our trend analysis approaches and sampling frequency recommendations to better inform public health departments how COVID-19 cases are changing, especially as rates of clinical testing continue to decline.

## Data Availability

All data are available through the Stanford Digital Repository at https://doi.org/10.25740/yg713sw8276.

## Acknowledgements

We thank the County of Santa Clara Public Health Department and the California Department of Public Health COVID-19 Wastewater Surveillance, Epidemiology and Data teams for their feedback about the trend analysis methods. Numerous people contributed to the collection of wastewater samples including Payak Sarkar, Noel Enoki, and Amy Wong.

## Author Contributions

**Elana M. G. Chan**: Conceptualization, Data Curation, Formal Analysis, Methodology, Software, Visualization, Writing (Original Draft Preparation, Reviewing & Editing); **Lauren C. Kennedy:** Conceptualization, Methodology, Writing (Original Draft Preparation, Reviewing & Editing); **Marlene K. Wolfe:** Conceptualization, Methodology, Writing (Reviewing & Editing); **Alexandria B. Boehm:** Conceptualization, Methodology, Writing (Original Draft Preparation, Reviewing & Editing), Supervision, Project Administration, Funding Acquisition.

## Data Availability

Wastewater data are publicly available through the Stanford Digital Repository (https://doi.org/10.25740/yg713sw8276).

## Supporting Information

**S1 Text. Supporting Information.**

