## Supplemental Information for "Identifying trends in SARS-CoV-2 RNA in wastewater to infer changing COVID-19 incidence: Effect of sampling frequency"

### Table of Contents

**1.0 SAN JOSÉ-SANTA CLARA REGIONAL WASTEWATER FACILITY SEWERSHED** 2

**2.0 TREND ANALYSIS METHODS** 3

2.1. Relative Strength Index 3

2.2. Percent Change 5

2.3. Mann-Kendall Trend Test 6

**3.0 REFERENCES** 8

### 1.0 SAN JOSÉ-SANTA CLARA REGIONAL WASTEWATER FACILITY SEWERSHED

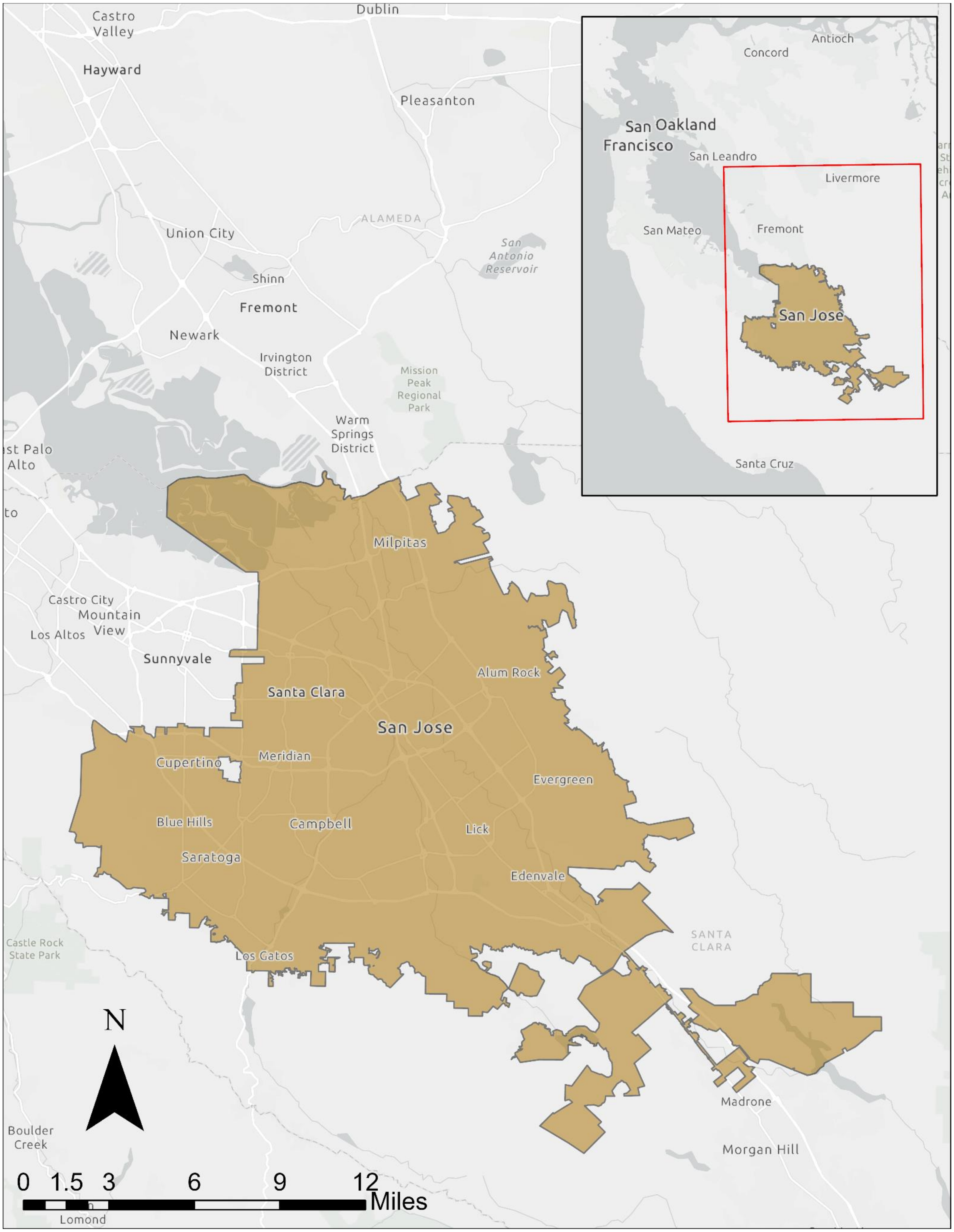


**Fig 1.** Map of the San José-Santa Clara Regional Wastewater Facility sewershed. Service layer credits: California State Parks, Esri, HERE, Garmin, FAO, NOAA, USGS, Bureau of Land Management, EPA, NPS, County of Santa Clara, SafeGraph, METI/NASA, USDA. Credit to Laura Roldan for creating the sewershed boundary layer

### 2.0 TREND ANALYSIS METHODS

#### 2.1. Relative Strength Index

Wilder's relative strength index (RSI) is a momentum indicator that measures the speed and direction of price movements and is calculated using the following equation [1]: $RSI = 100 - \frac{100}{1+RS}$, where $RS = \frac{average\ gain}{average\ loss}$. Using the standard 14-d look-back period, the first average gain (i.e., on day 14) is calculated as follows: $\sum \frac{gains\ over\ past\ 14\ days}{14}$. All subsequent average gains (i.e., day 15 and onwards) are calculated using a smoothing technique as follows: $\frac{previous\ average\ gain \times 13 + current\ gain}{14}$. The first and subsequent average losses are calculated in the same manner. Gains and losses on each day *t* are defined below where *p* refers to closing price. For this analysis, we used the 7-d moving average (MA) of N/PMMoV instead of closing prices as the input to the RSI calculation.

$\Delta_t = p_t - p_{t-1}$

when $\Delta_t > 0, gain_t = \Delta_t\ (loss_t = 0)$

when $\Delta_t < 0, loss_t = |\Delta_t|\ (gain_t = 0)$

Based on previously published work [1–3], we interpreted RSI values above 70, 80, and 90 as upward, likely upward, and very likely upward trends, respectively. We interpreted RSI values below 30, 20, and 10 as downward, likely downward, and very likely downward trends, respectively.

First, we explored using raw N/PMMoV values versus the MA of N/PMMoV as the input to the RSI equation (Fig 2). The two heatmaps below both use a 14-d look-back period, and the MA is specifically a 7-d, untrimmed, right-aligned MA. Using the MA of N/PMMoV versus raw N/PMMoV values as the input to the RSI equation provides more useful trend insight.

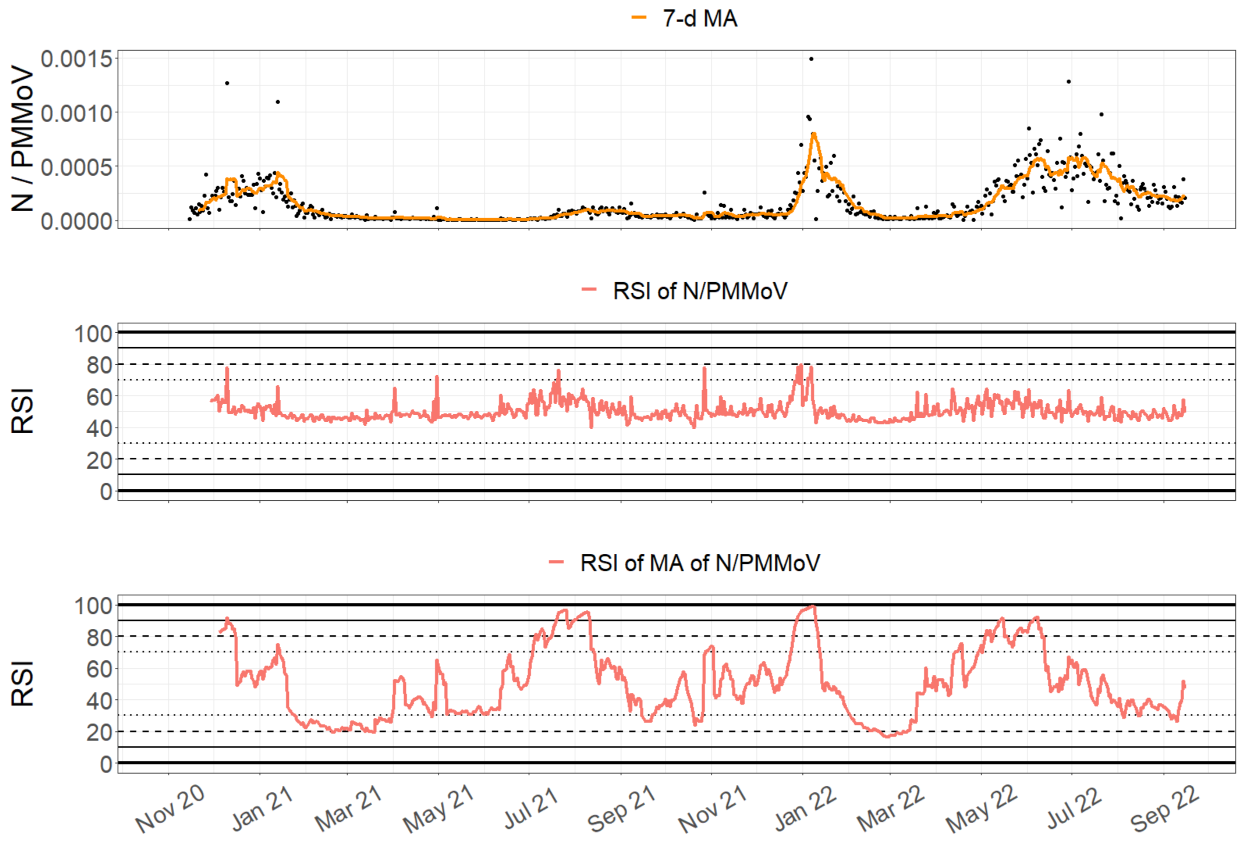


**Fig 2.** (Top) Daily N/PMMoV measurements with a 7-d moving average (MA). (Below) Daily RSI values (i) using raw N/PMMoV values as the input to the RSI calculation and (ii) using the 7-d MA of N/PMMoV as the input to the RSI calculation.

Next, we explored the effect of using different look-back periods on the RSI (7, 14, and 21 days) (Fig 3). The three heatmaps below all use the MA (7-d, untrimmed, right-aligned) of N/PMMoV as the input to the RSI equation. As the look-back period duration increases, the number of early signaling days in advance of major surges decreases. We ultimately decided to use the standard 14-d look-back period in our analysis which is similar to the look-back period used by the United States Centers for Disease Control (CDC) [4].

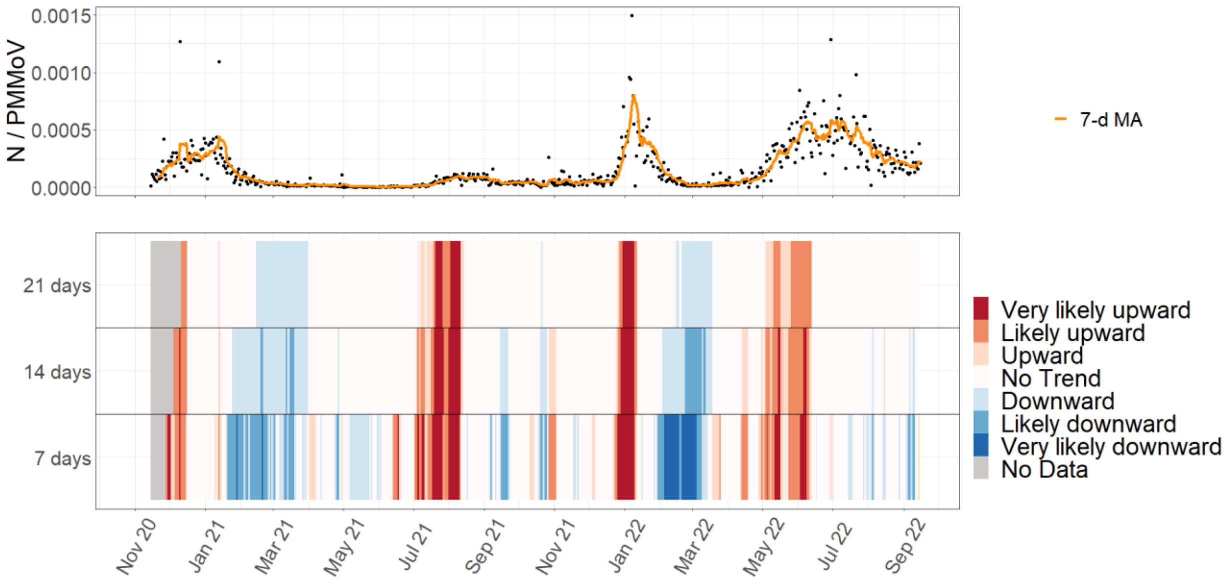


**Fig 3**. (Top) Daily N/PMMoV measurements with a 7-d moving average (MA). (Below) Daily RSI trend classifications (i) using a 21-d look-back period, (ii) using a 14-d look-back period, and (iii) using a 7-d look-back period. "No Data" indicates that not enough measurements were available yet to calculate an RSI value.

### 2.2. Percent Change

We used the following percent change (PC) formula from the United States Centers for Disease Control (CDC): $PC = \left(10^{slope} - 1\right) \times 100$, where *slope* is the slope from a least-squares linear regression of $\log_{10}$(N/PMMoV) versus day for the current day and previous 14 days (i.e., 15-d look-back period) [4]. For each slope estimate, we extracted the 90%, 95%, and 99% confidence intervals (CIs) to calculate the 90%, 95%, and 99% CIs for each PC estimate. We interpreted PC values greater than 0 for the 90%, 95%, and 99% upper CIs as upward, likely upward, and very likely upward trends, respectively. We interpreted PC values less than 0 for the 90%, 95%, and 99% lower CIs as downward, likely downward, and very likely downward trends, respectively. Below we explored different look-back periods for the slope parameter (7, 14, and 21 days) (Fig 4). We ultimately decided to use a 14-d look-back period for the slope in our analysis for consistency.

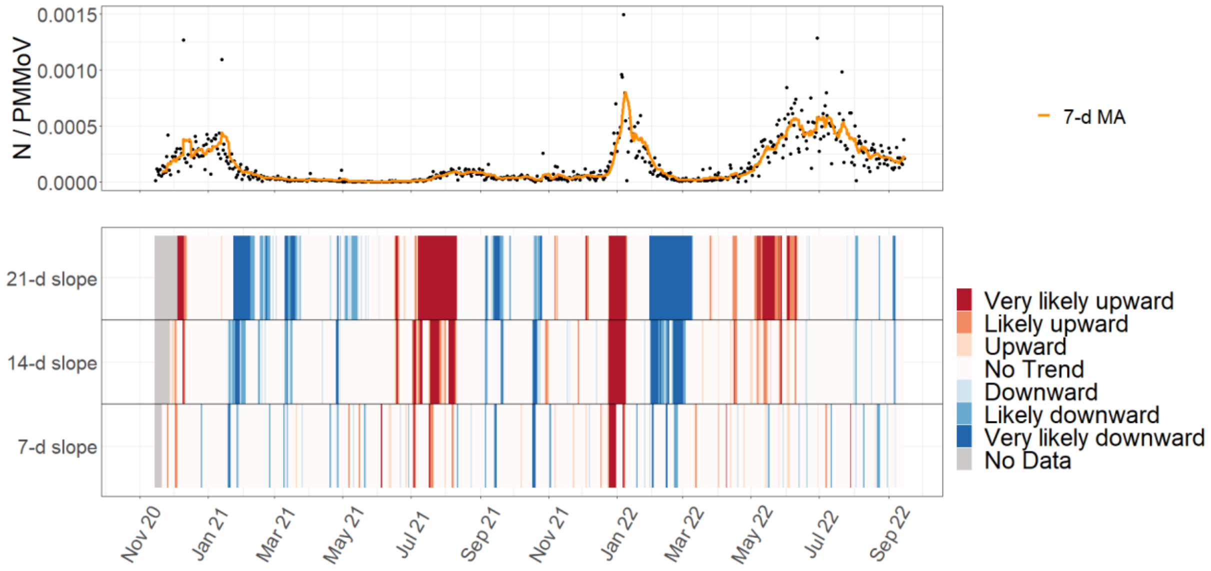


**Fig 4.** (Top) Daily N/PMMoV measurements with a 7-d moving average (MA). (Below) Daily percent change trend classifications using (i) a 21-d look-back period for the slope input, (ii) a 14-d look-back period for the slope input, and (ii) a 7-d look-back period for the slope input. "No Data" indicates that not enough measurements were available yet to calculate percent change.

#### 2.3. Mann-Kendall Trend Test

The Mann-Kendall (MK) trend test is a non-parametric test that evaluates whether there is a monotonic upward or downward trend in a time-series dataset [5–7]. We interpreted positive values of the test statistic, tau (τ), with $p \leq 0.1$, $p \leq 0.05$, and $p \leq 0.01$ as upward, likely upward, and very likely upward trends, respectively. We interpreted negative τ values with $p \leq 0.1$, $p \leq 0.05$, and $p \leq 0.01$ as downward, likely downward, and very likely downward trends, respectively. Below we explored different look-back periods over which to conduct the MK trends test (7, 14, and 21 days) (Fig 5). We ultimately decided to use a 14-d look-back period in our analysis for consistency.

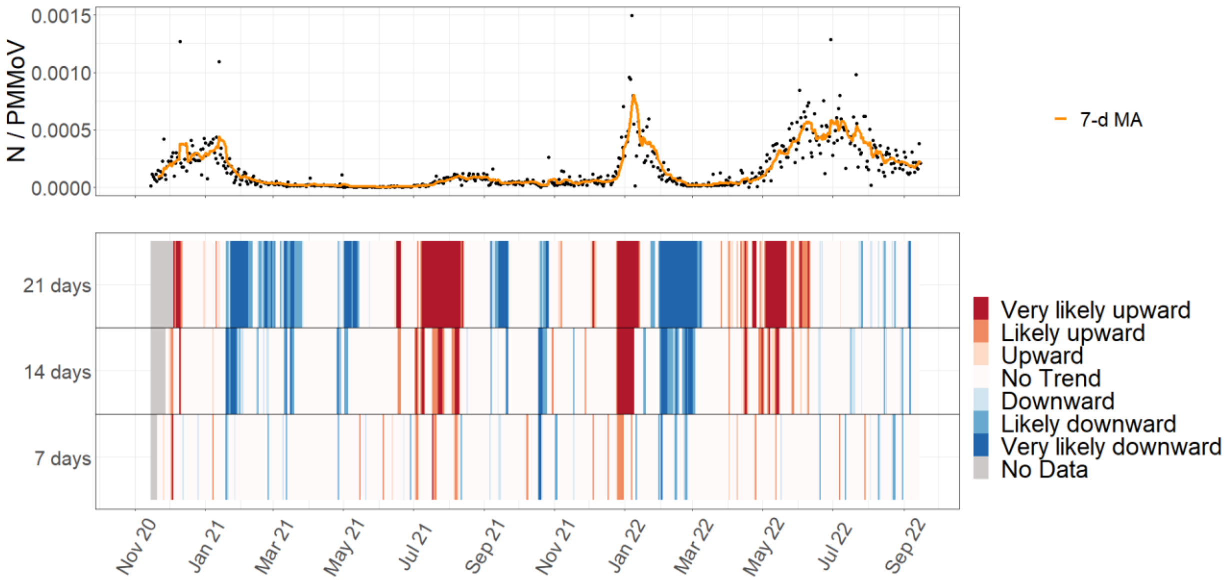


**Fig 5.** (Top) Daily N/PMMoV measurements with a 7-d moving average (MA). (Below) Daily Mann-Kendall trend classifications using (i) a 21-d look-back period, (ii) a 14-d look-back period, and (ii) a 7-d look-back period. “No Data” indicates that not enough measurements were available yet to conduct the Mann-Kendall trend test.
